# Causal Effects of Type 2 Diabetes and Glycemic Traits on Dementia and Stroke: A Mendelian Randomization Study including Imaging Endpoints

**DOI:** 10.1101/2025.06.11.25329399

**Authors:** Si Han, Elnaz Naderi, Kan Wang, Yuan Ma, Geert Jan Biessels, Fariba Ahmadizar

## Abstract

**Aims/hypothesis:** Type 2 diabetes mellitus (T2D) and glycemic dysregulation have been associated with cognitive decline and cerebrovascular disease, but causal relationships remain uncertain. We used Mendelian randomization (MR) to investigate the potential causal effects of T2D and glycemic traits, including hemoglobin A1c (HbA1c), fasting glucose (FG), fasting insulin (FI), and 2-hour glucose (2h-glu), on risk of dementia subtypes, stroke subtypes, and structural brain changes.

**Methods:** We conducted a two-sample MR analysis using summary statistics from large genome-wide association studies in European populations. Outcomes included all-cause dementia, Alzheimer’s disease (AD), vascular dementia (VaD), ischemic stroke, lacunar stroke, and five MRI-derived brain phenotypes (total brain volume, total white and grey matter volumes, hippocampal volume, and white matter hyperintensities (WMH)). The inverse-variance weighted method was the primary analysis; MR-Egger, weighted median, and Mendelian Randomization Pleiotropy RESidual Sum and Outlier (MR-PRESSO) were used for sensitivity analyses.

Multivariable MR adjusted for body mass index (BMI).

**Results:** Genetic liability to higher HbA1c was associated with increased AD risk (odds ratio (OR) = 1.35, 95% CI: 1.08-1.68) and lower hippocampal volume (β =-0.489, SE = 0.152). T2D was associated with increased risk of ischemic stroke (OR = 1.14, 95% CI: 1.11-1.17), lacunar stroke (OR = 1.15, 95% CI: 1.09-1.21), and lower grey matter volume (β =-0.032, SE = 0.011). FG, FI, and 2h-glu were also associated with stroke subtypes. These associations persisted after BMI adjustment. No consistent associations were observed with VaD or WMH.

**Conclusions**/**interpretation:** This study provides genetic evidence that HbA1c may causally contribute to AD and regional brain atrophy, while T2D and other glycemic traits are primarily linked to stroke risk. Incorporating brain MRI outcomes provides insight into distinct vascular and neurodegenerative pathways linking dysglycemia to brain health.

**Research in context:** *What is already known about this subject?:*

- Type 2 diabetes (T2D) and hyperglycemia have been associated with increased risk of dementia, particularly Alzheimer’s disease (AD), in observational studies.
- Observational designs are prone to confounding and reverse causality, limiting causal inference.
- Previous Mendelian randomization (MR) studies on glycemic traits and dementia risk have produced inconsistent results and rarely examined brain structural changes as intermediate outcomes.

*What is the key question?:*

- Do genetically predicted T2D and glycemic traits (hemoglobin A1c, fasting glucose, fasting insulin, 2-hour glucose) causally influence dementia risk, stroke subtypes, and neuroimaging biomarkers of brain structure?

*What are the new findings?:*

- Genetically predicted higher HbA1c levels were associated with increased risk of AD and with reductions in hippocampal and white matter volumes.
- T2D and other glycemic traits showed stronger associations with ischemic and small vessel stroke, suggesting vascular mechanisms.
- This study combines clinical and imaging outcomes to distinguish neurodegenerative from cerebrovascular pathways in diabetes-related brain changes.

*How might this impact clinical practice in the foreseeable future?:*

- The findings underscore the importance of optimizing long-term glycemic control even before the onset of T2D to reduce the risk of neurodegenerative and vascular brain pathology in individuals with diabetes.

## Introduction

Type 2 diabetes mellitus (T2D) and dysglycemia, including elevated hemoglobin A1c (HbA1c), fasting glucose (FG), and post-challenge glucose, represent major global health concerns, with over 530 million people affected worldwide, projected to rise to 783 million by 2045 (1). Beyond cardiovascular and renal complications, increasing evidence links glycemic dysregulation to adverse neurological outcomes, including dementia and stroke (2–4).

Dementia, including Alzheimer’s disease (AD) and vascular dementia (VaD), is projected to affect over 150 million people globally by 2050 (5). Identifying modifiable risk factors is a public health priority. Although observational studies suggest that T2D and related metabolic traits increase dementia risk (6–8), these associations are susceptible to confounding and reverse causation. Similarly, dysglycemia has been associated with ischemic and small vessel stroke (9, 10), yet causal inference remains limited (11, 12). Notably, the close interconnection between stroke and dementia necessitates their integrated consideration in both research and prevention. As highlighted in the Berlin Manifesto (37), these conditions share common risk factors and pathophysiology, with stroke nearly doubling the risk of subsequent dementia. Given its higher prevalence and greater potential for prevention, targeting stroke offers a compelling avenue to mitigate dementia risk at the population level. Mendelian randomization (MR) uses genetic variants as instrumental variables to strengthen causal inference and minimize bias (13). While previous MR studies, including our own systematic review and MR meta-analysis, have found no strong evidence for a causal effect of genetically predicted T2D on AD (OR = 1.02, 95% CI: 1.00-1.04) (14), the role of other glycemic traits (e.g., FG, HbA1c) on dementia, stroke subtypes, and structural brain magnetic resonance imaging (MRI) changes remain understudied.

To address these gaps, we conducted a two-sample MR study to investigate the potential causal effects of genetically predicted T2D and four glycemic traits, including HbA1c, FG, fasting insulin (FI), and 2-hour glucose (2h-glu), on dementia subtypes, stroke outcomes, and MRI-derived brain phenotypes. Our study aims to clarify the vascular and neurodegenerative pathways through which glycemic dysregulation may influence brain health by integrating clinical endpoints with neuroimaging markers.

## Methods

### Study design and data sources

Two-sample MR analyses were conducted to estimate the causal associations between T2D and glycemic traits (i.e., HbA1c, FG, FI, 2h-glu) with dementia outcomes (all-cause dementia, AD, VaD), stroke outcomes (ischemic stroke and lacunar stroke), and five brain MRI markers (normalized brain volume, normalized grey matter volume, normalized white matter volume, hippocampus volume, and WMH). Summary-level data were obtained from large genome-wide association studies (GWAS) conducted primarily in individuals of European ancestry (15–17). These datasets were selected based on sample size, methodological quality, and relevance to our research questions (see Table 1). The study followed the STROBE-MR guidelines for reporting (see *Supplementary Table S1*) (18). The study design flow can be found in Figure 1. For MR, three core assumptions must be fulfilled: (1) genetic instruments are strongly associated with the exposures; (2) they are not associated with confounders; and (3) they affect outcomes only through the exposures.

**Figure 1.**
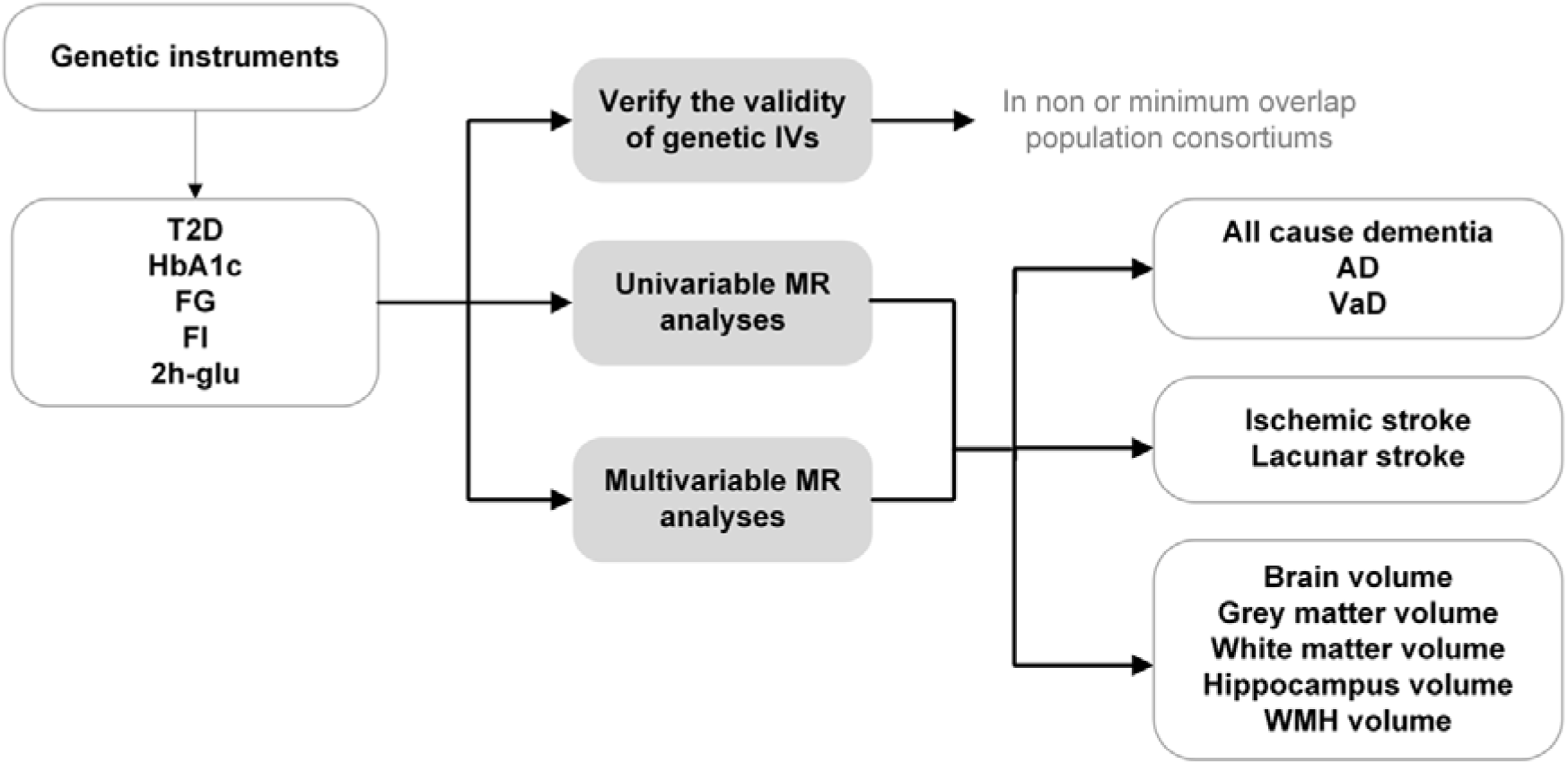
Overview of Study Design for Mendelian Randomization Analyses. T2D, type 2 diabetes; HbA1c, hemoglobin A1c; FG, fasting glucose; FI, fasting insulin; 2h-glu, 2-hour glucose; WMH: White Matter Hyperintensity volume; MR, Mendelian Randomization; IV, Instrumental Variable

**Table 1.** Characteristics of Exposure and Outcome GWAS Summary Statistics Used in the Mendelian Randomization Analyses.

| Phenotype | Consortium | Sample size (n-case/n-control) | Ancestry | Array Type | PMID |
| --- | --- | --- | --- | --- | --- |
| **T2D traits** |  |  |  |  |  |
| T2D | MVP, Diamante, PMB, MDCS, MedStar, PennCath | 148,726/965,732 | European | GWAS array and metabochip array | 32541925 |
|  | Mahajan 2018 | 81,412/370,832 | European | GWAS array | 29632382 |
|  | FinnGen | 32,469/183,185 | European | GWAS array | 36653562 |
|  | BioBank Japan | 36,614/155,150 | Asian | GWAS array | 30718926 |
| HbA1c | MAGIC | 149,006 | European | GWAS array and metabochip array | 34059833 |
| FG | MAGIC | 133,010 | European | GWAS array and metabochip array | 22885924 |
| FI | MAGIC | 108,557 | European | GWAS array and metabochip array | 22885924 |
| 2h-glu | MAGIC | 149,006 | European | GWAS array and metabochip array | 34059833 |
| **Dementia outcomes** |  |  |  |  |  |
| Dementia | FinnGen | 5,933 / 212,859 | European | GWAS array | 36653562 |
| AD | IGAP | 21,982/41,944 | European | GWAS array | 30820047 |
| VaD | FinnGen | 2,048/328,982 | European | GWAS array | 36653562 |
| Ischemic stroke | AFGen and others | 40,585/406,111 | European | GWAS array | 29531354 |
| Lacunar stroke | ISGC and others | 6,030 /248,929 | European | GWAS array | 33773637 |
| Brain volume | UKB Brain Imaging | 33,224 | European | GWAS array and metabochip array | 33875891 |
| Grey matter volume | UKB Brain Imaging | 33,224 | European | GWAS array and metabochip array | 33875891 |
| White matter volume | UKB Brain Imaging | 33,224 | European | GWAS array and metabochip array | 33875891 |
| Hippocampus volume | UKB Brain Imaging | 8,428 | European | GWAS array and metabochip array | 30305740 |
| WMH | UKB Brain Imaging | 32,114 | European | GWAS array and metabochip array | 33875891 |

*Abbreviations: n, number; T2D, type 2 diabetes; HbA1c, hemoglobin A1c; FG, fasting glucose; FI,* *fasting insulin; 2h-glu, 2-hour glucose; WMH: White Matter Hyperintensity volume; MVP, Million* *Veteran Program; PMB, Penn Medicine Biobank; MDCS, Malmo Diet and Cancer Study; MAGIC,* *Meta-Analyses of Glucose and Insulin-related traits Consortium; IGAP, International Genomics of* *Alzheimer's Project; ISGC, International Stroke Genetics Consortium; UKB, UK Biobank; GWAS,* *Genome-Wide Association Study*

### Instrumental variable selection and validation

To evaluate the core assumptions of MR outlined above, we took the following steps to select and validate the instrumental variables. Single-nucleotide polymorphisms (SNPs) associated with each exposure at genome-wide significance (p < 5×10^-8^) were selected and pruned for linkage disequilibrium (r² < 0.1 within a 1 Mb window). To minimize potential bias from horizontal pleiotropy, instrumental SNPs were screened against the GWAS Catalog for known associations with the outcomes of interest as well as traits representing potential alternative biological pathways. SNPs showing genome-wide significant associations with the outcomes and with traits likely to influence the outcomes independently of glycemic traits (e.g., inflammatory or vascular pathways) were excluded to reduce the risk of violating the exclusion restriction assumptions. Steiger filtering was subsequently applied to remove variants that explained more variance in the outcome than in the exposure, thereby reducing the risk of reverse causality. All retained SNPs exhibited F-statistics greater than 10, indicating adequate instrument strength. Validation analyses using independent GWAS datasets for the same traits (19–22) confirmed the consistency and directionality of associations, supporting the robustness of the selected instruments. Details of SNP selection, clumping, Steiger filtering, and validation across independent GWAS datasets are provided in *Supplementary Material S1*.

### Outcome definitions and GWAS sources

#### Dementia outcome data

For AD, we used summary statistics from the International Genomics of Alzheimer’s Project (IGAP), a large two-stage GWAS meta-analysis conducted in individuals of European ancestry (23). The stage 1 analysis, used here, included 17,008 clinically diagnosed AD cases and 37,154 controls from four major consortia: the Alzheimer’s Disease Genetics Consortium (ADGC), European Alzheimer’s Disease Initiative (EADI), and others, with genotyping and imputation data on over 7 million SNPs.

For vascular and all-cause dementia, outcome data were obtained from the FinnGen consortium (Freeze 8), which integrates genetic data with nationwide health registry records in a genetically homogeneous Finnish population (20). Dementia diagnoses were defined using ICD-10 codes. We included 7,284 all-cause dementia cases (209,487 controls) and 881 VaD cases (211,508 controls). These large-scale GWAS datasets are well-suited for MR analyses due to their phenotypic specificity, population consistency, and statistical power.

#### Stroke outcome data

We used summary-level data from two large GWAS to evaluate the causal effects of T2D and glycemic traits on stroke outcomes. For ischemic stroke, we obtained summary data from the MEGASTROKE consortium, which conducted a multi-ancestry GWAS meta-analysis including 40,585 stroke cases and 406,111 controls of European ancestry. Analyses were stratified by stroke subtype, and all MR analyses in the present study were restricted to participants of European descent to reduce population stratification (24).

For lacunar stroke, we used data from the International Stroke Genetics Consortium, which pooled newly recruited MRI-confirmed cases from the UK DNA Lacunar Stroke Studies with existing GWAS datasets. The final meta-analysis included 7,338 cases and 254,798 controls, including a subset of 2,987 cases matched to 29,540 controls with imaging-confirmed diagnosis. The use of MRI-confirmed lacunar stroke cases increases phenotype specificity and enhances the validity of MR estimates. Both datasets represent the largest available genetic resources for stroke subtypes and are well-suited for causal inference analyses (25).

#### Brain Imaging Phenotypes

We utilized GWAS summary statistics for five structural brain MRI traits from the UK Biobank brain imaging extension, which includes high-resolution MRI data from approximately 40,000 participants of European ancestry. Imaging-derived phenotypes (IDPs) were generated using standardized image processing pipelines, and all volumetric traits were normalized for head size (26).

The five outcomes included were: total brain volume (normalized for head size), reflecting the overall size of the brain parenchyma and serving as a global marker of brain atrophy; Grey matter volume (normalized for head size), which captures the volume of cortical and subcortical neuronal tissue; White matter volume (normalized for head size), representing the myelinated axonal tracts and structural connectivity; Hippocampal volume, a region critical to memory formation and one of the earliest affected in AD; and white matter hyperintensity (WMH), a radiological marker of cerebral small vessel disease and age-related brain pathology. WMH was log-transformed due to its right-skewed distribution. These phenotypes were selected to capture global and regional brain atrophy, neurodegeneration (hippocampus), and small vessel disease burden (WMH). The use of these large-scale, population-based imaging GWASs enables a robust investigation of the genetic determinants of brain structure and their potential causal relationships with metabolic traits (27, 28).

### Statistical methods

To ensure consistent effect allele orientation across exposure and outcome summary statistics, all SNPs were harmonized to reflect the same reference allele.

Palindromic SNPs (A/T or C/G) were resolved using allele frequency data, where available; ambiguous variants and those with strand incompatibility were excluded. When primary SNPs were unavailable in outcome datasets, proxies in high linkage disequilibrium (r² > 0.8) were used.

SNPs with a minor allele frequency (MAF) < 0.01 were excluded to minimize potential bias from rare alleles. Steiger filtering was applied to ensure instrument validity to exclude SNPs that explained more variance in the outcome than in the exposure, which suggests potential reverse causality.

The primary MR analysis under a fixed-effects model used the inverse-variance weighted (IVW) method with effect estimates scaled per one standard deviation (1-SD) increase for continuous traits or per unit increase in genetic liability for binary. Sensitivity analyses included MR-Egger regression (testing for directional pleiotropy via the intercept), Cochran’s Q statistic (heterogeneity), MR-PRESSO (outlier detection and correction), and the weighted median estimator (robust with up to 50% invalid instruments). Additional robustness checks included the simple and weighted mode-based estimators.

### Multivariable Mendelian Randomization (MVMR)

To assess whether the observed associations between glycemic traits and brain outcomes were independent of adiposity, we performed multivariable Mendelian randomization (MVMR) analyses adjusting for genetically predicted body mass index (BMI). We included only exposures significantly associated with outcomes in the univariable MR analyses. Independent genome-wide significant SNPs for each exposure (e.g., T2D, HbA1c) and BMI were harmonized across datasets. IVW was used to estimate the direct effect of each glycemic trait on brain outcomes, adjusting for BMI.

All analyses were performed using the *TwoSampleMR* (v0.5.10) package in RStudio (version 2024.04; Posit, PBC, Boston, MA, USA). All P values were two-sided. The Bonferroni correction was applied to account for multiple testing across outcomes and associations with a P value < 0.05 were considered statistically significant. Nominal P values < 0.05, not surviving correction, were interpreted as suggestive.

## Results

### Instrumental variable selection and validation

We selected strong, independent SNPs for T2D and glycemic traits, excluding those associated with study outcomes and alternative pathways. Specifically, rs1260326 (T2D and FI), associated with ischemic stroke and chronic inflammatory diseases, and rs1483121 (FG), associated with AD and BMI, were excluded to reduce potential pleiotropic bias. All instruments exceeded the F-statistic threshold of 10, indicating strong relevance. To further ensure validity, we conducted validation using independent GWAS datasets, which confirmed consistent associations in the expected directions. These findings support the robustness of the instruments and their suitability for MR analyses. Full details are provided in *Supplementary Material S2*, *Supplementary Table S2,* and *Supplementary Figure S1*.

### T2D and glycemic traits: associations with dementia outcomes

The results of the IVW analyses on the relationship between T2D, glycemic traits, and dementia outcomes are shown in Figure 2.

**Figure 2.**
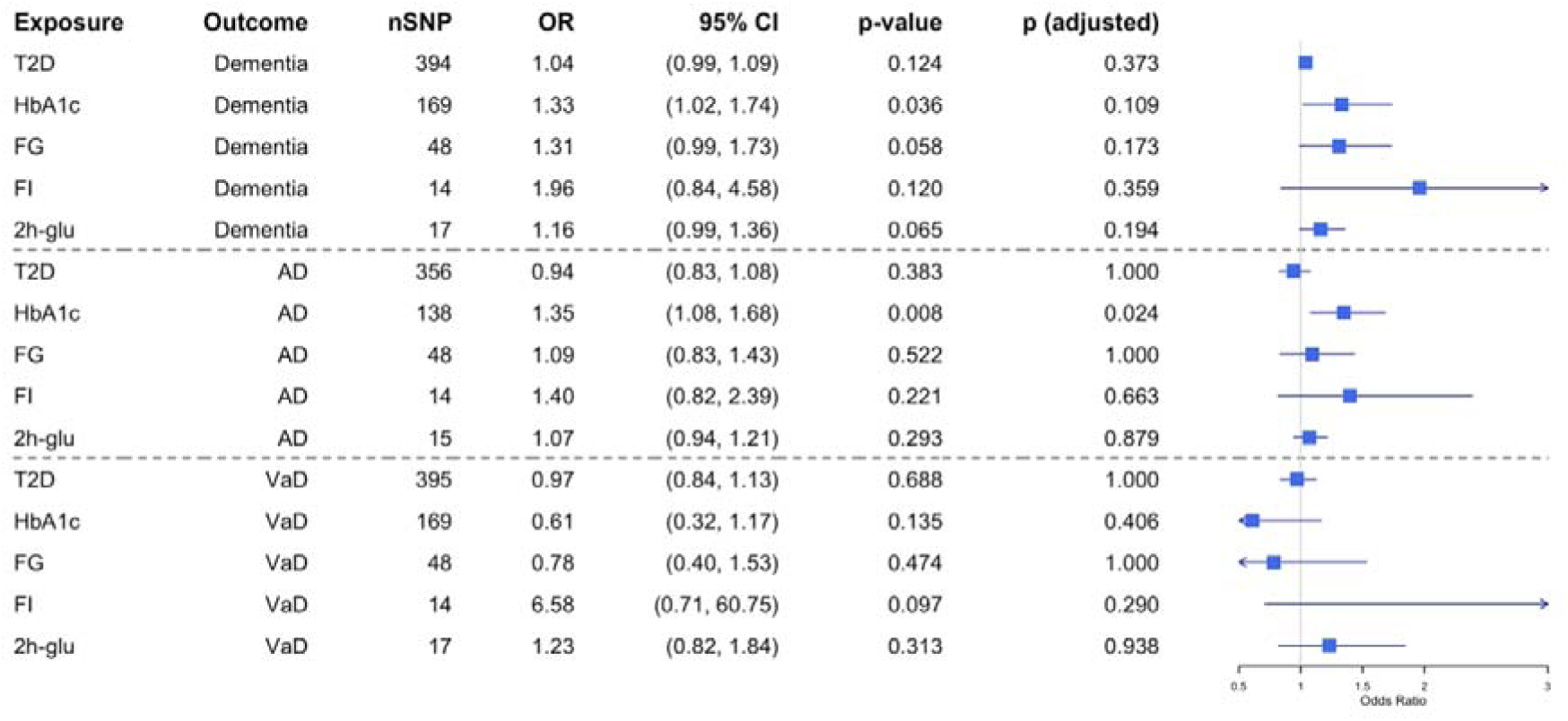
Mendelian Randomization Estimates for the Association Between T2D and Glycemic Traits with Dementia-Related Outcomes Using the Inverse-Variance Weighted Method T2D, type 2 diabetes; HbA1c, hemoglobin A1c; FG, fasting glucose; FI, fasting insulin; 2h-glu, 2-hour glucose; AD, Alzheimer’s disease; VaD, vascular dementia Odds ratios are shown for genetically predicted T2D, HbA1, FG, FI, and 2h-glu with dementia, AD, and VaD.

For all-cause dementia, none of the T2D and glycemic traits showed a robust causal association in IVW analyses. HbA1c showed a suggestive association (OR = 1.33, 95% CI: 1.02-1.74, p = 0.036), but this did not remain significant after Bonferroni correction. Sensitivity analyses (e.g., weighted median) suggested possible associations between FG and 2h-glu, but IVW estimates remained null. No evidence of pleiotropy or outliers was detected.

For AD, genetically predicted HbA1c was significantly associated with increased risk (OR = 1.35, 95% CI: 1.08-1.68, p = 0.008), surviving correction for multiple testing. No significant associations were observed for T2D, HbA1c, FG, FI, and 2h-glu. Some heterogeneity was observed for T2D and FG, but MR-Egger and MR-PRESSO tests did not indicate directional pleiotropy or distortion of effect estimates.

For VaD, none of the glycemic traits showed significant associations.

Although MR-PRESSO identified outlier variants in the T2D model, their removal did not meaningfully change the effect estimates or null findings. Across all exposures, tests for horizontal pleiotropy indicated no significant bias, and results from multiple sensitivity analyses consistently supported the robustness of null associations.

Full results, including pleiotropy and outlier diagnostics, are provided in *Supplementary Tables S3 and S4*.

### T2D and glycemic traits: associations with stroke outcomes

The results of MR analyses on associations between T2D, glycemic traits and ischemic or lacunar stroke are presented in Figure 3.

**Figure 3.**
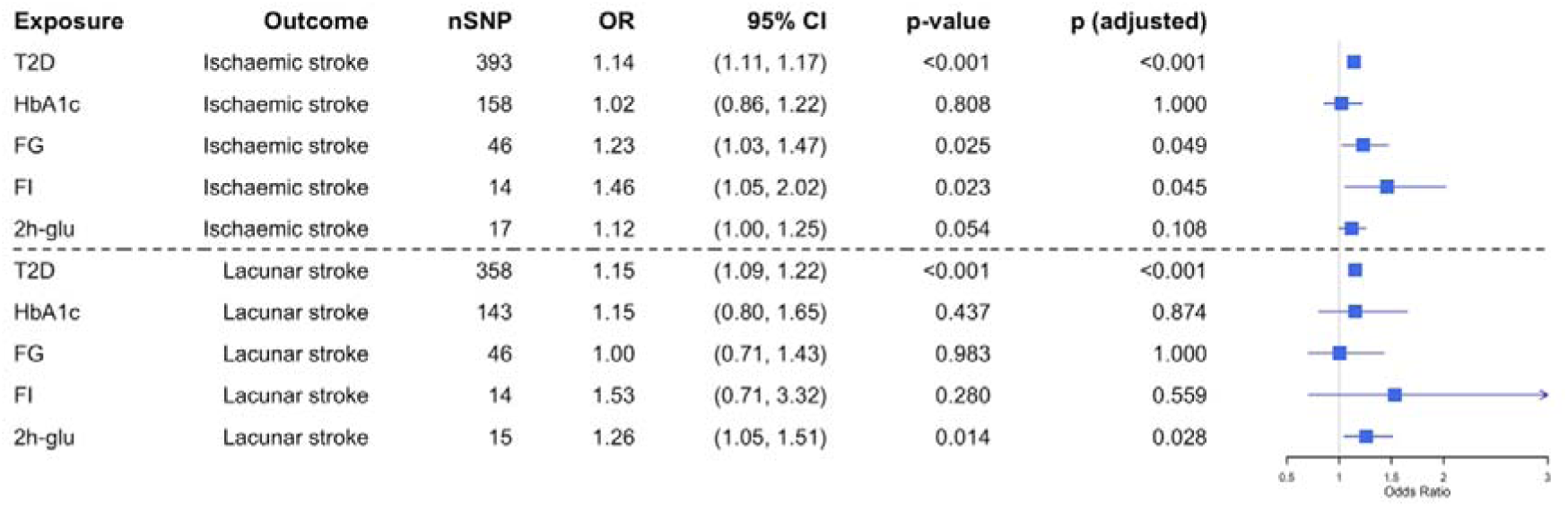
Mendelian Randomization Estimates for the Association Between T2D and Glycemic Traits with Stroke Subtypes Using the Inverse-Variance Weighted Method T2D, type 2 diabetes; HbA1c, hemoglobin A1c; FG, fasting glucose; FI, fasting insulin; 2h-glu, 2-hour glucose Odds ratios are shown for genetically predicted T2D, HbA1, FG, FI, and 2h-glu with two types of strokes.

**Figure 4.**
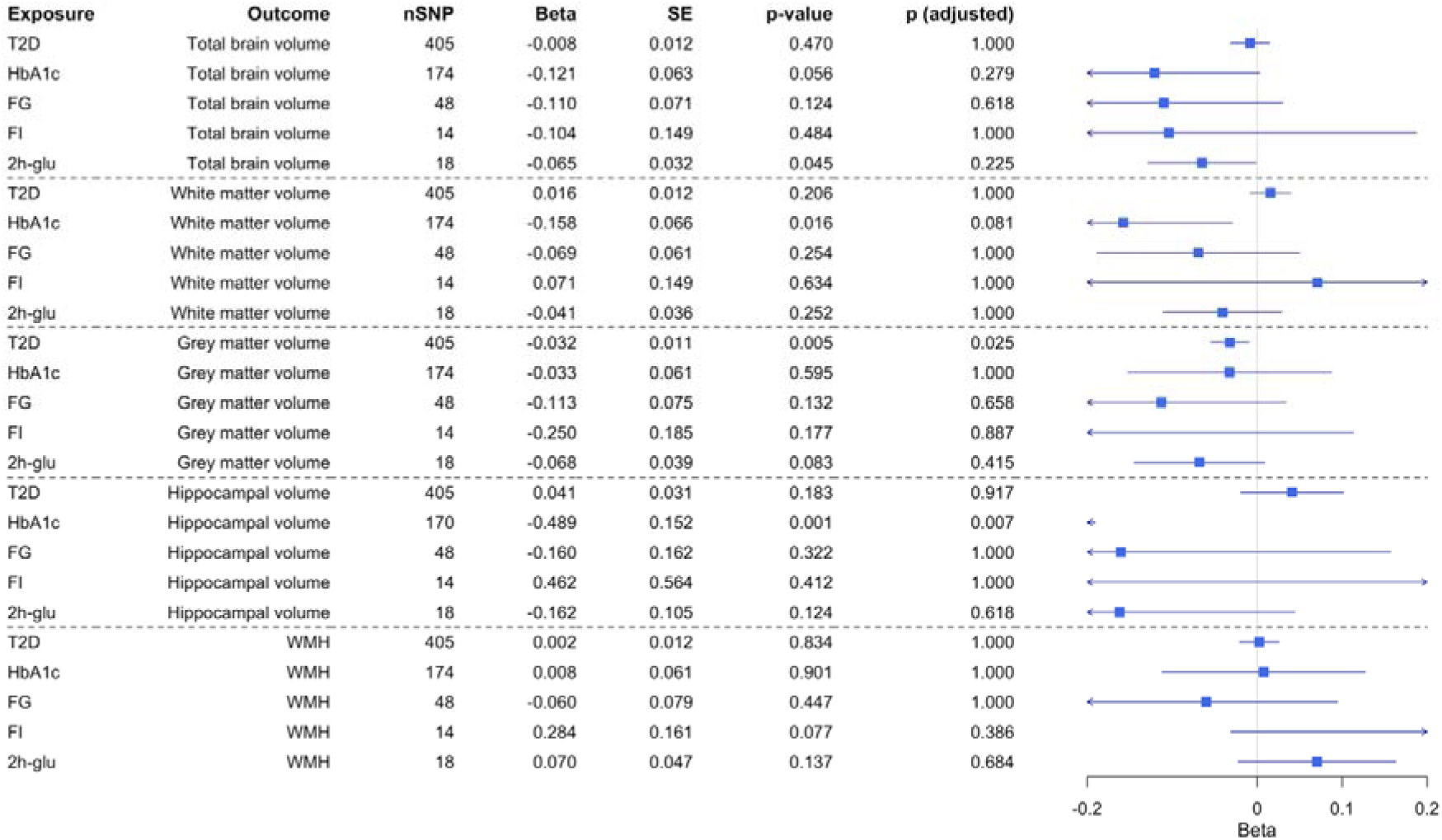
Mendelian Randomization Estimates for the Association Between T2D and Glycemic Traits with Brain MRI markers Using the Inverse-Variance Weighted Method T2D, type 2 diabetes; HbA1c, hemoglobin A1c; FG, fasting glucose; FI, fasting insulin; 2h-glu, 2-hour glucose; WMH, white matter hyperintensities Odds ratios are shown for genetically predicted T2D, HbA1, FG, FI, and 2h-glu with neuroimaging markers.

T2D was significantly associated with both ischemic stroke (IVW OR = 1.14, 95% CI: 1.11-1.17, p = 5.05 × 10^-27^) and lacunar stroke (IVW OR = 1.15, 95% CI: 1.09-1.22, p = 1.67 × 10^-7^), with results remaining significant after Bonferroni correction (Figure 3). Sensitivity analyses, including the MR-Egger and weighted median, yielded directionally consistent effects. For ischemic stroke, substantial heterogeneity was observed (Q = 501.55, degrees of freedom (df) = 392, *p* = 1.47 × 10^-4^), but no evidence of directional pleiotropy was found (MR-Egger intercept *p* = 0.159). MR-PRESSO identified four outlier SNPs, and exclusion did not meaningfully alter the effect estimate (*p* = 2.29 × 10^-26^). No heterogeneity or outliers were detected for lacunar stroke.

HbA1c was not significantly associated with either stroke subtype (Figure 3). For ischemic stroke, substantial heterogeneity was detected (Q = 356.05, df = 157, *p* = 1.71 × 10^-17^), though pleiotropy tests were negative. MR-PRESSO identified four outliers, but their removal did not change the inference. For lacunar stroke, IVW estimates were null, while MR-Egger (*p* = 0.045) and weighted median (*p* = 0.052) suggested a borderline association that remained insignificant after multiple testing correction. MR-PRESSO identified three outliers; the corrected estimate remained non-significant (*p* = 0.211), and pleiotropy was borderline (*p* = 0.060).

Genetically predicted FG was significantly associated with ischemic stroke in the IVW analysis (OR = 1.23, 95% CI: 1.03-1.47, *p* = 0.025), which remained statistically significant after Bonferroni correction (adjusted *p* = 0.049), as shown in Figure 3. However, MR-PRESSO identified three outliers, and removal attenuated the association. No association was found with lacunar stroke. FI was significantly associated with ischemic stroke, which remained robust after Bonferroni correction (OR = 1.46, 95% CI: 1.05-2.02, *p* = 0.023), robust across sensitivity models and without evidence of pleiotropy or outliers. No associations were observed with lacunar stroke.

2h-glu was not associated with ischemic stroke. However, it was significantly associated with lacunar stroke (OR 1.26; 95% CI 1.05, 1.51; p = 0.014), which remained significant after Bonferroni correction (Figure 3). No heterogeneity, pleiotropy, or outliers were detected.

The full results, including sensitivity analyses, can be found in *Supplementary Tables S5 and S6*.

### T2D and glycemic traits with brain imaging outcomes

T2D was significantly associated with reduced grey matter volume (β =-0.032, SE = 0.011, *p* = 0.005), and this association remained significant after Bonferroni correction and MR-PRESSO outlier adjustment (Figure 3). No associations were found for total brain volume, white matter volume, hippocampal volume, or WMH (all *p* > 0.20). While moderate heterogeneity was observed in some models, there was no evidence of directional pleiotropy, supporting a potential region-specific effect of T2D on cortical grey matter.

HbA1c showed a significant inverse association with hippocampal volume (β =-0.49, SE = 0.152, *p* = 0.001), which remained significant after Bonferroni correction. A suggestive association with white matter volume (β =-0.158; p = 0.016), which strengthened after outlier correction (*p* = 0.006) but did not retain significance after correction for multiple testing. No significant associations were observed for other imaging traits, and sensitivity analyses confirmed the robustness of results.

FG, FI, and 2h-glu showed no significant associations with MRI outcomes after correction. A nominal association between 2h-glu and total brain volume (β = - 0.065; p = 0.045) remained insignificant after adjustment. Across all three traits, there was no consistent evidence of pleiotropy, and MR-PRESSO corrections did not materially alter the null findings.

The full results from sensitivity models and pleiotropy assessments are reported in the *Supplementary Tables S7 and S8*.

### Multivariable Mendelian Randomization (MVMR)

For T2D, associations with both ischemic stroke (OR 1.14; 95% CI 1.12, 1.17; p = 1.27 × 10^-27^) and lacunar stroke (OR 1.15; 95% CI 1.09, 1.21; p = 1.68 × 10^-7^) remained robust. BMI was not significantly associated with either stroke subtype (ischemic stroke: OR 1.02; 95% CI 0.93, 1.10; p = 0.688; lacunar stroke: OR 1.03; 95% CI 0.83, 1.28; p = 0.784), indicating that BMI does not likely confound these associations.

For grey matter volume, the inverse association with T2D observed in univariable MR also persisted in the MVMR model (β =-0.032; SE = 0.011; p = 0.005). The effect of BMI in this model was insignificant (β = 0.053; SE = 0.047; p = 0.261), again suggesting a BMI-independent effect.

The association between HbA1c and AD remained statistically significant after adjustment (OR = 1.31, 95% CI: 1.03-1.67, *p* = 0.027). BMI was not significantly associated with AD risk (OR = 0.77, 95% CI: 0.42-1.32, *p* = 0.390), suggesting that the HbA1c-AD association is unlikely to be confounded by BMI.

MVMR analyses for HbA1c-white matter volume and 2h-glu-lacunar stroke could not be performed due to limited instrument availability.

## Discussion

This is the first MR study to integrate clinical outcomes and quantitative brain MRI data to investigate T2D and glycemic traits’ causal effects on dementia and ischemic stroke and related brain MRI features.

Here we show that genetically predicted T2D is causally associated with an increased risk of ischemic and lacunar stroke, consistent with and extending previous MR findings (11, 29, 30). These associations reinforce the role of vascular mechanisms - including atherosclerosis, microvascular dysfunction, and chronic inflammation – as key pathways linking metabolic dysregulation to cerebrovascular events (31, 32). The specific association with lacunar stroke, a subtype of small vessel disease, highlights the role of microangiopathy and supports previous findings of cerebral small vessel damage in diabetes (33, 34).

In contrast, we found no strong evidence for a direct causal effect of T2D on all-cause dementia, AD, or VaD aligning with prior MR meta-analyses (14). This suggests that while T2D is a clear risk factor for vascular events, its role in neurodegeneration may be mediated through other glycemic mechanisms or comorbidities.

Beyond clinical endpoints, we also examined the association between T2D and structural brain changes. Genetically predicted T2D was significantly associated with reduced grey matter volume, while no associations were observed for total brain volume, white matter volume, hippocampal volume, or WMH. These findings suggest a region-specific vulnerability of cortical grey matter to the effects of T2D, possibly reflecting early neurodegenerative or vascular injury processes.

Among individual glycemic traits, genetically predicted HbA1c showed a distinct pattern of associations. Elevated HbA1c levels were linked to increased risk of AD, while other glycemic traits (FG, FI, and 2h-glu) were primarily associated with stroke subtypes, further implicating glycemic traits in vascular risk beyond T2D diagnosis.

In imaging analyses, higher HbA1c was also associated with reduced hippocampal and white matter volumes. These structural changes are consistent with early neurodegenerative and vascular injury and may precede overt clinical symptoms. The inverse association with hippocampal volume is particularly notable given the hippocampus’s early involvement in AD. This supports clinical evidence linking long-term hyperglycemia to cognitive decline (35) and may reflect neurodegenerative processes driven by chronic hyperglycemia, oxidative stress, and accumulation of advanced glycation end-products (AGEs) (36). Likewise, reduced white matter volume may signal early axonal damage, another hallmark of AD’s pathology (37, 38).

To further probe potential confounding by adiposity, we conducted MVMR analyses adjusting for genetically predicted BMI. The associations between T2D and both ischemic and lacunar stroke and T2D and reduced grey matter volume remained significant after adjustment, suggesting independence from obesity-related pathways. Similarly, the HbA1c-AD association persisted with only slight attenuation. However, multivariable MR could not be conducted for other significant univariable findings (e.g., HbA1c-white matter volume; 2h-glu-lacunar stroke) due to limited instrument overlap, and these associations should be interpreted cautiously.

Although we did not observe consistent associations with VaD or WMH, this may reflect limitations in clinical diagnosis within population-based studies. VaD is often underdiagnosed, and many dementia cases, particularly in older adults, involve mixed pathologies that blur distinctions between subtypes (39, 40). WMH, while a known marker of small vessel disease, may not directly correlate with clinically diagnosed VaD due to these overlaps. Given these complexities, focusing on all-cause dementia as the primary outcome may provide a more comprehensive measure of disease burden in this context.

A key strength of this study is the use of MR analysis to minimize confounding biases and reverse causation. It also applies multivariable MR to account for BMI and includes quantitative brain MRI outcomes to explore mechanistic pathways beyond clinical endpoints. Using multiple sensitivity methods and validation across independent GWAS strengthens the robustness of our findings.

Nevertheless, several limitations should be considered when interpreting these findings. First, our analyses were restricted to individuals of European ancestry, which limits the generalizability of results to non-European populations. Second, power was limited for some outcomes, particularly for VaD, due to small sample sizes. Third, although we applied Steiger filtering, reverse causality cannot be entirely excluded for imaging outcomes. Fourth, instrument strength varied across exposures through rigorous instrumental selection and validation processes.

Notably, the genetic instrument for fasting insulin included only 15 SNPs, raising the possibility of weak instrument bias and reduced power to detect causal effects.

Finally, as the HbA1c GWAS was conducted in non-diabetic individuals, the findings may not fully reflect the risk in people with established T2D.

### Clinical implications and future studies

Our findings indicate that HbA1c may contribute to neurodegeneration independently of diabetes, while T2D and related traits are causally linked to stroke risk. This highlights the importance of early glycemic control to reduce vascular and neurodegenerative brain damage. Monitoring HbA1c, even in non-diabetic individuals, may help identify those at risk for AD or brain atrophy. Future studies should assess whether improving glycemic control in midlife can modify brain structure or cognitive decline. Longitudinal imaging and intervention studies in diverse populations are needed to define actionable glycemic thresholds and inform precision prevention strategies.

## Abbreviations

T2D: Type 2 diabetes
HbA1c: Hemoglobin A1c
FG: Fasting glucose
FI: Fasting insulin
2h-glu: 2-hour glucose
GWAS: Genome-wide association study
IVW: Inverse weighted variance
AD: Alzheimer’s disease
VAD: Vascular dementia
WMH: White matter hyperintensity
MR: Mendelian Randomization
MAF: Minor allele frequency
LD: Linkage disequilibrium

## Data availability

The GWAS summary statistics used in this study are publicly available from the following sources: https://www.ebi.ac.uk/gwas/ (accessed on 1st Dec 2024). Data on glycemic traits have been contributed by MAGIC investigators and have been downloaded from www.magicinvestigators.org.

## Authors’ relationships and activities

The authors declare that no relationships or activities could appear to have influenced the submitted work.

## Contribution statement

S.H., F.A. and G.J.B. conceived the idea presented. S.H. conducted all statistical analyses under the supervision of F.A. and G.J.B.. S.H. drafted the manuscript, with F.A. providing critical revisions and feedback. All authors have reviewed and approved the final version of the manuscript.

## Data Availability

All data produced in the present study are available upon reasonable request to the authors

