## Supplemental Materials for "Causal Effects of Type 2 Diabetes and Glycemic Traits on Dementia and Stroke: A Mendelian Randomization Study including Imaging Endpoints"

Supplementary Table S1. STROBE-MR checklist of recommended items to address in reports of Mendelian randomization studies

Supplementary Material S1. Instrumental variable selection and validation methods

Supplementary Material S2. Instrumental variable selection and validation results

Supplementary Table S2. Summary of genetic variant harmonization between exposure and outcome datasets

Supplementary Figure S1. Forest plot of instrumental variables validation for T2D and glycemic traits

Supplementary Table S3. Mendelian randomization analysis for the causal associations between T2D and clinically diagnosed dementia outcomes based on five methods

Supplementary Table S4. Mendelian randomization analysis for the causal associations between glycemic traits and clinically diagnosed dementia outcomes based on five methods

Supplementary Table S5. Mendelian randomization analysis for the causal associations between T2D and clinically diagnosed stroke outcomes based on five methods

Supplementary Table S6. Mendelian randomization analysis for the causal associations between glycemic traits and clinically diagnosed stroke outcomes based on five methods

Supplementary Table S7. Mendelian randomization analysis for the causal associations between T2D and brain MRI markers based on five methods

Supplementary Table S8. Mendelian randomization analysis for the causal associations between glycemic traits and brain MRI markers based on five methods

**Supplementary Table S1.** **STROBE-MR checklist of recommended items to address in reports of Mendelian randomization studies**

| **Item No.** | **Section** | **Checklist item** | **Page No.** | **Relevant text from manuscript** |
| --- | --- | --- | --- | --- |
| 1 | **TITLE and ABSTRACT** | Indicate Mendelian randomization (MR) as the study’s design in the title and/or the abstract if that is a main purpose of the study | 2 | Abstract |
|  | **INTRODUCTION** |  |  |  |
| 2 | **Background** | Explain the scientific background and rationale for the reported study. What is the exposure? Is a potential causal relationship between exposure and outcome plausible? Justify why MR is a helpful method to address the study question | 5 | Introduction |
| 3 | **Objectives** | State specific objectives clearly, including pre-specified causal hypotheses (if any). State that MR is a method that, under specific assumptions, intends to estimate causal effects | 5 | Introduction |
|  | **METHODS** |  |  |  |
| 4 | **Study design and data sources** | Present key elements of the study design early in the article. Consider including a table listing sources of data for all phases of the study. For each data source contributing to the analysis, describe the following: |  |  |
|  | a) | Setting: Describe the study design and the underlying population, if possible. Describe the setting, locations, and relevant dates, including periods of recruitment, exposure, follow-up, and data collection, when available. | 6 | Study design and data sources |
|  | b) | Participants: Give the eligibility criteria, and the sources and methods of selection of participants. Report the sample size, and whether any power or sample size calculations were carried out prior to the main analysis | 6 | Study design and data sources |
|  | c) | Describe measurement, quality control and selection of genetic variants | 7 | Instrumental variable selection and validation |
|  | d) | For each exposure, outcome, and other relevant variables, describe methods of assessment and diagnostic criteria for diseases | 7 | Outcome definitions and GWAS sources |
|  | e) | Provide details of ethics committee approval and participant informed consent, if relevant |  |  |
| 5 | **Assumptions** | Explicitly state the three core IV assumptions for the main analysis (relevance, independence and exclusion restriction) as well assumptions for any additional or sensitivity analysis | 7 | Study design and data sources |
| 6 | **Statistical methods: main analysis** | Describe statistical methods and statistics used |  |  |
|  | a) | Describe how quantitative variables were handled in the analyses (i.e., scale, units, model) | 9-10 | Statistical methods |
|  | b) | Describe how genetic variants were handled in the analyses and, if applicable, how their weights were selected | 9-10 | Statistical methods |
|  | c) | Describe the MR estimator (e.g. two-stage least squares, Wald ratio) and related statistics. Detail the included covariates and, in case of two-sample MR, whether the same covariate set was used for adjustment in the two samples | 9-10 | Statistical methods |
|  | d) | Explain how missing data were addressed |  |  |
|  | e) | If applicable, indicate how multiple testing was addressed | 9-10 | Statistical methods |
| 7 | **Assessment of assumptions** | Describe any methods or prior knowledge used to assess the assumptions or justify their validity | 9-10 | Statistical methods |
| 8 | **Sensitivity analyses and additional analyses** | Describe any sensitivity analyses or additional analyses performed (e.g. comparison of effect estimates from different approaches, independent replication, bias analytic techniques, validation of instruments, simulations) | 9-10 | Statistical methods |
| 9 | **Software and pre-registration** |  |  |  |
|  | a) | Name statistical software and package(s), including version and settings used | 9-10 | Statistical methods |
|  | b) | State whether the study protocol and details were pre-registered (as well as when and where) | 9-10 | Statistical methods |
|  | **RESULTS** |  |  |  |
| 10 | **Descriptive data** |  |  |  |
|  | a) | Report the numbers of individuals at each stage of included studies and reasons for exclusion. Consider use of a flow diagram |  | Supplementary Material S1 and S2 |
|  | b) | Report summary statistics for phenotypic exposure(s), outcome(s), and other relevant variables (e.g. means, SDs, proportions) | 20 | Table 1 |
|  | c) | If the data sources include meta-analyses of previous studies, provide the assessments of heterogeneity across these studies |  |  |
|  | d) | For two-sample MR:  i.  Provide justification of the similarity of the genetic variant-exposure associations between the exposure and outcome samples  ii.  Provide information on the number of individuals who overlap between the exposure and outcome studies |  | Supplementary Material S1 and S2 |
| 11 | **Main results** |  |  |  |
|  | a) | Report the associations between genetic variant and exposure, and between genetic variant and outcome, preferably on an interpretable scale | 11-15 | Results |
|  | b) | Report MR estimates of the relationship between exposure and outcome, and the measures of uncertainty from the MR analysis, on an interpretable scale, such as odds ratio or relative risk per SD difference | 11-15 | Results |
|  | c) | If relevant, consider translating estimates of relative risk into absolute risk for a meaningful time period | 11-15 | Results |
|  | d) | Consider plots to visualize results (e.g. forest plot, scatterplot of associations between genetic variants and outcome versus between genetic variants and exposure) | 11-15 | Results |
| 12 | **Assessment of assumptions** |  |  |  |
|  | a) | Report the assessment of the validity of the assumptions | 11-15 | Results |
|  | b) | Report any additional statistics (e.g., assessments of heterogeneity across genetic variants, such as *I^2^*, Q statistic or E-value) | 11-15 | Results |
| 13 | **Sensitivity analyses and additional analyses** |  |  |  |
|  | a) | Report any sensitivity analyses to assess the robustness of the main results to violations of the assumptions | 11-15 | Results |
|  | b) | Report results from other sensitivity analyses or additional analyses | 11-15 | Results |
|  | c) | Report any assessment of direction of causal relationship (e.g., bidirectional MR) |  |  |
|  | d) | When relevant, report and compare with estimates from non-MR analyses | 15-19 | Discussion |
|  | e) | Consider additional plots to visualize results (e.g., leave-one-out analyses) | 11-15 | Results |
|  | **DISCUSSION** |  |  |  |
| 14 | **Key results** | Summarize key results with reference to study objectives | 15-19 | Discussion |
| 15 | **Limitations** | Discuss limitations of the study, taking into account the validity of the IV assumptions, other sources of potential bias, and imprecision. Discuss both direction and magnitude of any potential bias and any efforts to address them | 15-19 | Discussion |
| 16 | **Interpretation** |  |  |  |
|  | a) | Meaning: Give a cautious overall interpretation of results in the context of their limitations and in comparison with other studies | 15-19 | Discussion |
|  | b) | Mechanism: Discuss underlying biological mechanisms that could drive a potential causal relationship between the investigated exposure and the outcome, and whether the gene-environment equivalence assumption is reasonable. Use causal language carefully, clarifying that IV estimates may provide causal effects only under certain assumptions | 15-19 | Discussion |
|  | c) | Clinical relevance: Discuss whether the results have clinical or public policy relevance, and to what extent they inform effect sizes of possible interventions | 15-19 | Discussion |
| 17 | **Generalizability** | Discuss the generalizability of the study results (a) to other populations, (b) across other exposure periods/timings, and (c) across other levels of exposure | 15-19 | Discussion |
|  | **OTHER INFORMATION** |  |  |  |
| 18 | **Funding** | Describe sources of funding and the role of funders in the present study and, if applicable, sources of funding for the databases and original study or studies on which the present study is based | 19 | Funding |
| 19 | **Data and data sharing** | Provide the data used to perform all analyses or report where and how the data can be accessed, and reference these sources in the article. Provide the statistical code needed to reproduce the results in the article, or report whether the code is publicly accessible and if so, where | 19 | Data availability |
| 20 | **Conflicts of Interest** | All authors should declare all potential conflicts of interest | 19 | Authors’ relationships and activities |

This checklist is copyrighted by the Equator Network under the Creative Commons Attribution 3.0 Unported (CC BY 3.0) license.

**Supplementary Material S1. Instrumental variable selection and validation methods**

***Instrument selection for exposures***

Single nucleotide polymorphisms (SNPs) associated with each exposure at genome-wide significance (p < 5×10^-8^) were selected as instrumental variables (IVs). For type 2 diabetes (T2D), we used a meta-analysis of approximately 1.4 million participants, of which 148,726 European cases and 965,732 European controls in the Million Veteran Program, DIAMANTE, Malmo Diet and Cancer Study, and other consortiums, which identified 318 novel risk loci for T2D and related vascular outcomes (1). For glycemic traits, including hemoglobin A1c (HbA1c), fasting glucose (FG), fasting insulin (FI), and 2-hour glucose (2h-glu), IVs were sourced from Meta-Analyses of Glucose and Insulin-related traits Consortium (MAGIC). We prioritized SNPs from non-BMI-adjusted models to avoid potential collider bias, except or 2h-glu for which only BMI-adjusted summary statistics were available.

We applied clumping (r² < 0.1, window size 1 Mb) using the 1000 Genomes European reference panel to ensure independence of instruments. F-statistics were calculated for each SNP using the formula:

$$F=\frac{\left( N-2 \right)\cdot R^{2}}{1-R^{2}}$$

where R was derived from the SNP’s effect size and standard error. SNPs with F ≤ 10 were excluded. To reduce horizontal pleiotropy, we screened each variant using GWAS Catalog to check for associations with potential confounders or outcomes.

***Instrumental variables validation***

For glycemic traits, additional MR analyses were conducted using outcome data from the UK Biobank for HbA1c (N ≈ 407,000) and from MAGIC meta-analyses covering FG, FI, and 2h-glu in up to 281,416 individuals (approximately 30% non-European ancestry). This approach enabled cross-validation of MR findings for glycemic traits using independent outcome sources and diverse study populations, increasing confidence in the robustness of our causal inferences. To assess the robustness and generalizability of our MR findings, we repeated key analyses using the same genetic instruments across independent genome-wide association study (GWAS) outcome datasets.

For T2D, three well-powered and ancestrally diverse datasets were used (1) the GWAS by Mahajan et al. (2018), , which combined exome array and GWAS data from 81,412 T2D cases and 370,832 controls across multiple populations (2); (2) the FinnGen consortium, , which included 49,303 T2D cases identified from national registries in a Finnish population-based cohort (3); and (3) the Biobank Japan (BBJ), which comprised 36,614 T2D cases and 155,150 controls of Japanese ancestry and identified 88 genome-wide significant loci, including 30 novel associations (4). These complementary datasets allowed us to examine the consistency of causal estimates across populations with different genetic backgrounds, phenotyping methods, and healthcare systems.

For glycemic traits, additional MR analyses were conducted using outcome data from the UK Biobank for HbA1c (approximately 407,000) (5) and from MAGIC meta-analyses covering FG, FI, and 2h-glu in up to 281,416 individuals (approximately 30% non-European ancestry). This approach enabled cross-validation of MR findings for glycemic traits using independent outcome sources and diverse study populations, increasing confidence in the robustness of our causal inferences.

**Supplementary Material S2. Instrumental variable selection and validation results**

***Instrumental variable selection***

During the filtering in GWAS Catalog, one SNP for T2D (rs1260326), one SNP for FI (rs1260326) were associated with ischemic stroke, and with chronic inflammatory diseases. One SNP (rs1483121) for FG was associated with Alzheimer’s disease (AD) and body mass index (BMI), which all were excluded. After clumping and excluding the outcome related SNPs, we used the remaining 416 SNPs for T2D, 178 SNPs for HbA1c, 50 SNPs for FG, 15 SNPs for FI, and 18 for 2h-glu as the instrument in the harmonization and Steiger filter analysis to check if each SNP explains more variance in the exposure than in the outcome to rule out reverse causation. All SNPs exceeded the conventional threshold of F>10, indicating that the genetic variants explain a significant portion of the variance in the exposure variable.

***Instrumental variables validation***

To validate the selected instrumental variables for each glycemic trait, we performed MR analyses using independent GWAS datasets of the same trait as proxy outcomes. For T2D, the IVs were tested against three independent GWAS datasets (OR 2.31, 95% CI 2.23-2.39 for Mahajan 2018 GWAS; OR 2.39, 95% CI 2.30-2.48 for FinnGen consortium; OR 2.29, 95% CI 2.16-2.42 for Biobank Japan), all of which demonstrated robust and statistically significant associations in the expected direction. Similar validation analyses were performed for HbA1c, FG, FI, and 2h-glu, each showing consistent causal estimates across alternative datasets. These findings confirm the strength and consistency of the instruments, supporting their use in subsequent MR analyses.

**Supplementary Table S2. Summary of genetic variant harmonization between exposure and outcome datasets**

| Exposure | Outcome | Initial SNPs | Matched SNPs | Palindromic Removed | Strand Mismatch | Steiger pass | Final SNPs |
| --- | --- | --- | --- | --- | --- | --- | --- |
| **Dementia outcomes** | | | | | | | |
| T2D | Dementia | 416 | 408 | 14 | 0 | 394 | 394 |
|  | AD | 416 | 409 | 53 | 0 | 356 | 356 |
|  | VaD | 416 | 409 | 14 | 0 | 395 | 395 |
| HbA1c | Dementia | 178 | 174 | 5 | 0 | 174 | 169 |
|  | AD | 178 | 167 | 28 | 0 | 166 | 138 |
|  | VaD | 178 | 174 | 5 | 0 | 174 | 169 |
| FG | Dementia | 50 | 50 | 2 | 0 | 50 | 48 |
|  | AD | 50 | 50 | 2 | 0 | 50 | 48 |
|  | VaD | 50 | 50 | 2 | 0 | 50 | 48 |
| FI | Dementia | 15 | 15 | 1 | 0 | 15 | 14 |
|  | AD | 15 | 15 | 1 | 0 | 15 | 14 |
|  | VaD | 15 | 15 | 1 | 0 | 15 | 14 |
| 2h-glu | Dementia | 18 | 18 | 1 | 0 | 18 | 17 |
|  | AD | 18 | 18 | 3 | 0 | 18 | 15 |
|  | VaD | 18 | 18 | 1 | 0 | 18 | 17 |
| **Stroke outcomes** | | | | | | | |
| T2D | Ischemic stroke | 416 | 409 | 16 | 0 | 409 | 393 |
|  | Lacunar stroke | 416 | 412 | 54 | 0 | 412 | 358 |
| HbA1c | Ischemic stroke | 178 | 158 | 0 | 0 | 158 | 158 |
|  | Lacunar stroke | 178 | 143 | 0 | 0 | 143 | 143 |
| FG | Ischemic stroke | 50 | 46 | 0 | 0 | 46 | 46 |
|  | Lacunar stroke | 50 | 46 | 0 | 0 | 46 | 46 |
| FI | Ischemic stroke | 15 | 15 | 1 | 0 | 15 | 14 |
|  | Lacunar stroke | 15 | 15 | 1 | 0 | 15 | 14 |
| 2h-glu | Ischemic stroke | 18 | 17 | 0 | 0 | 17 | 17 |
|  | Lacunar stroke | 18 | 18 | 3 | 0 | 18 | 15 |
| **Brain volumes** | | | | | | | |
| T2D | Brain volume | 416 | 417 | 11 | 1 | 417 | 405 |
|  | Grey matter volume | 416 | 417 | 11 | 1 | 417 | 405 |
|  | White matter volume | 416 | 417 | 11 | 1 | 417 | 405 |
|  | Hippocampus volume | 416 | 416 | 10 | 1 | 416 | 405 |
|  | WMH | 416 | 417 | 11 | 1 | 417 | 405 |
| HbA1c | Brain volume | 178 | 178 | 4 | 0 | 178 | 174 |
|  | Grey matter volume | 178 | 178 | 4 | 0 | 178 | 174 |
|  | White matter volume | 178 | 178 | 4 | 0 | 178 | 174 |
|  | Hippocampus volume | 178 | 174 | 4 | 0 | 174 | 170 |
|  | WMH | 178 | 178 | 4 | 0 | 178 | 174 |
| FG | Brain volume | 50 | 50 | 2 | 0 | 50 | 48 |
|  | Grey matter volume | 50 | 50 | 2 | 0 | 50 | 48 |
|  | White matter volume | 50 | 50 | 2 | 0 | 50 | 48 |
|  | Hippocampus volume | 50 | 50 | 2 | 0 | 50 | 48 |
|  | WMH | 50 | 50 | 2 | 0 | 50 | 48 |
| FI | Brain volume | 15 | 15 | 1 | 0 | 15 | 14 |
|  | Grey matter volume | 15 | 15 | 1 | 0 | 15 | 14 |
|  | White matter volume | 15 | 15 | 1 | 0 | 15 | 14 |
|  | Hippocampus volume | 15 | 15 | 1 | 0 | 15 | 14 |
|  | WMH | 15 | 15 | 1 | 0 | 15 | 14 |
| 2h-glu | Brain volume | 18 | 18 | 0 | 0 | 18 | 18 |
|  | Grey matter volume | 18 | 18 | 0 | 0 | 18 | 18 |
|  | White matter volume | 18 | 18 | 0 | 0 | 18 | 18 |
|  | Hippocampus volume | 18 | 18 | 0 | 0 | 18 | 18 |
|  | WMH | 18 | 18 | 0 | 0 | 18 | 18 |

***Abbreviations:*** *T2D: Type 2 diabetes; HbA1c: Hemoglobin A1c; FG: Fasting glucose; FI: Fasting insulin; 2h-glu: 2-hour glucose; AD: Alzheimer's disease; VaD: Vascular dementia; WMH: White matter hyperintensities.*

**Supplementary Figure S1. Forest plot of instrumental variables validation for T2D and glycemic traits**

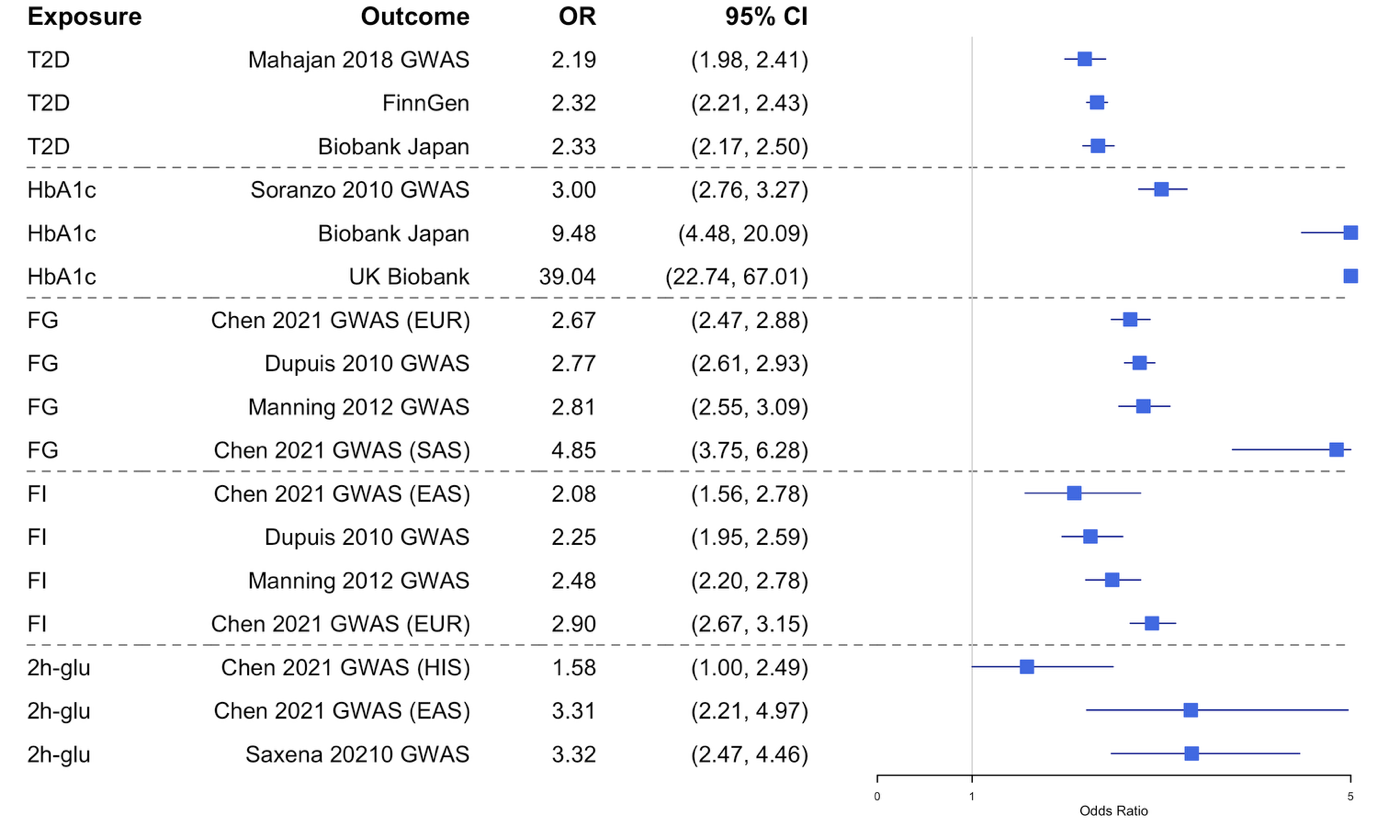

***Abbreviations:*** *T2D, type 2 diabetes; HbA1c: Hemoglobin A1c; FG: Fasting glucose; FI: Fasting insulin; 2h-glu: 2-hour glucose; OR: odds ratio; CI: confidence interval.*

**Supplementary Table S3. Mendelian randomization analysis for the causal associations between T2D and clinically diagnosed dementia outcomes based on five methods**

| Outcomes | Method | nSNP | p-value | OR | LCI | UCI |
| --- | --- | --- | --- | --- | --- | --- |
| Dementia | MR Egger | 394 | 0.70 | 1.02 | 0.92 | 1.14 |
|  | Weighted median | 394 | 0.12 | 1.08 | 0.98 | 1.19 |
|  | Inverse variance weighted | 394 | 0.12 | 1.04 | 0.99 | 1.09 |
|  | Simple mode | 394 | 0.79 | 0.97 | 0.78 | 1.21 |
|  | Weighted mode | 394 | 0.36 | 1.06 | 0.94 | 1.20 |
| AD | MR Egger | 356 | 0.24 | 0.85 | 0.64 | 1.12 |
|  | Weighted median | 356 | 1.00 | 1.00 | 0.93 | 1.08 |
|  | Inverse variance weighted | 356 | 0.38 | 0.94 | 0.83 | 1.08 |
|  | Simple mode | 356 | 0.26 | 1.12 | 0.92 | 1.37 |
|  | Weighted mode | 356 | 0.89 | 1.01 | 0.93 | 1.09 |
| VaD | MR Egger | 395 | 0.85 | 0.97 | 0.70 | 1.34 |
|  | Weighted median | 395 | 0.63 | 1.07 | 0.82 | 1.38 |
|  | Inverse variance weighted | 395 | 0.69 | 0.97 | 0.84 | 1.13 |
|  | Simple mode | 395 | 0.54 | 0.82 | 0.44 | 1.53 |
|  | Weighted mode | 395 | 0.88 | 1.02 | 0.76 | 1.39 |

***Abbreviations:*** *n: number; T2D: type 2 diabetes; AD: Alzheimer's disease; VaD: Vascular dementia; SNP: Single-nucleotide polymorphism; OR: Odds ratio; LCI: Lower confidence interval; UCI: Upper confidence interval.*

**Supplementary Table S4. Mendelian randomization analysis for the causal associations between glycemic traits and clinically diagnosed dementia outcomes based on five methods**

| Exposure | Outcomes | Method | nSNP | p-value | OR | LCI | UCI |
| --- | --- | --- | --- | --- | --- | --- | --- |
| HbA1c | Dementia | MR Egger | 169 | 0.12 | 1.55 | 0.89 | 2.69 |
|  |  | Weighted median | 169 | 0.75 | 1.07 | 0.71 | 1.61 |
|  |  | Inverse variance weighted | 169 | 0.04 | 1.33 | 1.02 | 1.74 |
|  |  | Simple mode | 169 | 0.51 | 0.72 | 0.27 | 1.90 |
|  |  | Weighted mode | 169 | 0.87 | 0.94 | 0.47 | 1.89 |
|  | AD | MR Egger | 138 | 0.05 | 1.53 | 1.00 | 2.34 |
|  |  | Weighted median | 138 | 0.05 | 1.42 | 1.00 | 2.03 |
|  |  | Inverse variance weighted | 138 | 0.01 | 1.35 | 1.08 | 1.68 |
|  |  | Simple mode | 138 | 0.73 | 1.12 | 0.59 | 2.14 |
|  |  | Weighted mode | 138 | 0.13 | 1.32 | 0.92 | 1.88 |
|  | VaD | MR Egger | 169 | 0.98 | 0.98 | 0.26 | 3.75 |
|  |  | Weighted median | 169 | 0.46 | 0.67 | 0.24 | 1.92 |
|  |  | Inverse variance weighted | 169 | 0.14 | 0.61 | 0.32 | 1.17 |
|  |  | Simple mode | 169 | 0.85 | 1.26 | 0.11 | 14.44 |
|  |  | Weighted mode | 169 | 0.41 | 0.43 | 0.06 | 3.15 |
| FG | Dementia | MR Egger | 48 | 0.07 | 1.74 | 0.97 | 3.10 |
|  |  | Weighted median | 48 | 0.03 | 1.56 | 1.04 | 2.33 |
|  |  | Inverse variance weighted | 48 | 0.06 | 1.31 | 0.99 | 1.73 |
|  |  | Simple mode | 48 | 0.98 | 0.99 | 0.43 | 2.25 |
|  |  | Weighted mode | 48 | 0.03 | 1.81 | 1.08 | 3.05 |
|  | AD | MR Egger | 48 | 0.15 | 1.52 | 0.87 | 2.67 |
|  |  | Weighted median | 48 | 0.06 | 1.36 | 0.99 | 1.87 |
|  |  | Inverse variance weighted | 48 | 0.52 | 1.09 | 0.83 | 1.43 |
|  |  | Simple mode | 48 | 0.69 | 1.14 | 0.60 | 2.18 |
|  |  | Weighted mode | 48 | 0.17 | 1.25 | 0.91 | 1.72 |
|  | VaD | MR Egger | 48 | 0.48 | 0.60 | 0.15 | 2.45 |
|  |  | Weighted median | 48 | 0.84 | 0.90 | 0.30 | 2.71 |
|  |  | Inverse variance weighted | 48 | 0.47 | 0.78 | 0.40 | 1.53 |
|  |  | Simple mode | 48 | 0.64 | 1.56 | 0.25 | 9.63 |
|  |  | Weighted mode | 48 | 0.99 | 1.01 | 0.30 | 3.42 |
| FI | Dementia | MR Egger | 14 | 0.15 | 0.04 | 0.00 | 2.35 |
|  |  | Weighted median | 14 | 0.08 | 2.49 | 0.89 | 6.97 |
|  |  | Inverse variance weighted | 14 | 0.12 | 1.96 | 0.84 | 4.58 |
|  |  | Simple mode | 14 | 0.47 | 2.14 | 0.29 | 15.95 |
|  |  | Weighted mode | 14 | 0.63 | 1.65 | 0.23 | 11.80 |
|  | AD | MR Egger | 14 | 0.07 | 22.14 | 1.10 | 443.58 |
|  |  | Weighted median | 14 | 0.13 | 1.74 | 0.85 | 3.54 |
|  |  | Inverse variance weighted | 14 | 0.22 | 1.40 | 0.82 | 2.39 |
|  |  | Simple mode | 14 | 0.24 | 2.09 | 0.64 | 6.80 |
|  |  | Weighted mode | 14 | 0.17 | 2.16 | 0.76 | 6.15 |
|  | VaD | MR Egger | 14 | 0.72 | 0.10 | 0.00 | 21,086.17 |
|  |  | Weighted median | 14 | 0.04 | 16.65 | 1.12 | 248.18 |
|  |  | Inverse variance weighted | 14 | 0.10 | 6.58 | 0.71 | 60.75 |
|  |  | Simple mode | 14 | 0.13 | 89.43 | 0.41 | 19,340.17 |
|  |  | Weighted mode | 14 | 0.15 | 56.56 | 0.31 | 10,311.91 |
| 2h-glu | Dementia | MR Egger | 17 | 0.08 | 1.57 | 0.98 | 2.53 |
|  |  | Weighted median | 17 | 0.03 | 1.27 | 1.02 | 1.57 |
|  |  | Inverse variance weighted | 17 | 0.06 | 1.16 | 0.99 | 1.36 |
|  |  | Simple mode | 17 | 0.19 | 1.30 | 0.89 | 1.90 |
|  |  | Weighted mode | 17 | 0.12 | 1.33 | 0.95 | 1.87 |
|  | AD | MR Egger | 15 | 0.95 | 1.01 | 0.70 | 1.48 |
|  |  | Weighted median | 15 | 0.18 | 1.13 | 0.94 | 1.35 |
|  |  | Inverse variance weighted | 15 | 0.29 | 1.07 | 0.94 | 1.21 |
|  |  | Simple mode | 15 | 0.15 | 1.31 | 0.93 | 1.84 |
|  |  | Weighted mode | 15 | 0.16 | 1.27 | 0.93 | 1.75 |
|  | VaD | MR Egger | 17 | 0.56 | 1.46 | 0.43 | 4.95 |
|  |  | Weighted median | 17 | 0.65 | 1.14 | 0.66 | 1.96 |
|  |  | Inverse variance weighted | 17 | 0.31 | 1.23 | 0.82 | 1.84 |
|  |  | Simple mode | 17 | 0.98 | 1.01 | 0.41 | 2.54 |
|  |  | Weighted mode | 17 | 0.87 | 1.07 | 0.47 | 2.44 |

***Abbreviations:*** *n: number; HbA1c: Hemoglobin A1c; FG: Fasting glucose; FI: Fasting insulin; 2h-glu: 2-hour glucose; AD: Alzheimer's disease; VaD: Vascular dementia; SNP: Single-nucleotide polymorphism; OR: Odds ratio; LCI: Lower confidence interval; UCI: Upper confidence interval.*

**Supplementary Table S5. Mendelian randomization analysis for the causal associations between T2D and clinically diagnosed stroke outcomes based on five methods**

| Outcomes | Method | nSNP | p-value | OR | LCI | UCI |
| --- | --- | --- | --- | --- | --- | --- |
| Ischemic stroke | MR Egger | 393 | 0.00 | 1.10 | 1.05 | 1.16 |
|  | Weighted median | 393 | 0.00 | 1.14 | 1.09 | 1.19 |
|  | Inverse variance weighted | 393 | 0.00 | 1.14 | 1.11 | 1.17 |
|  | Simple mode | 393 | 0.03 | 1.12 | 1.01 | 1.25 |
|  | Weighted mode | 393 | 0.00 | 1.12 | 1.06 | 1.18 |
| Lacunar stroke | MR Egger | 358 | 0.02 | 1.14 | 1.02 | 1.27 |
|  | Weighted median | 358 | 0.01 | 1.16 | 1.03 | 1.31 |
|  | Inverse variance weighted | 358 | 0.00 | 1.15 | 1.09 | 1.22 |
|  | Simple mode | 358 | 0.00 | 1.63 | 1.24 | 2.13 |
|  | Weighted mode | 358 | 0.07 | 1.12 | 0.99 | 1.26 |

***Abbreviations:*** *n: number; T2D: type 2 diabetes; SNP: Single-nucleotide polymorphism; OR: Odds ratio; LCI: Lower confidence interval; UCI: Upper confidence interval.*

**Supplementary Table S6. Mendelian randomization analysis for the causal associations between glycemic traits and clinically diagnosed stroke outcomes based on five methods**

| Exposure | Outcomes | Method | nSNP | p-value | OR | LCI | UCI |
| --- | --- | --- | --- | --- | --- | --- | --- |
| HbA1c | Ischemic stroke | MR Egger | 158 | 0.67 | 0.92 | 0.64 | 1.33 |
|  |  | Weighted median | 158 | 0.13 | 0.85 | 0.70 | 1.05 |
|  |  | Inverse variance weighted | 158 | 0.81 | 1.02 | 0.86 | 1.22 |
|  |  | Simple mode | 158 | 0.24 | 0.78 | 0.52 | 1.18 |
|  |  | Weighted mode | 158 | 0.08 | 0.80 | 0.63 | 1.03 |
|  | Lacunar stroke | MR Egger | 143 | 0.04 | 2.02 | 1.02 | 3.98 |
|  |  | Weighted median | 143 | 0.29 | 1.29 | 0.81 | 2.07 |
|  |  | Inverse variance weighted | 143 | 0.44 | 1.15 | 0.80 | 1.65 |
|  |  | Simple mode | 143 | 0.85 | 0.90 | 0.29 | 2.81 |
|  |  | Weighted mode | 143 | 0.05 | 1.98 | 1.00 | 3.93 |
| FG | Ischemic stroke | MR Egger | 46 | 0.66 | 0.92 | 0.62 | 1.35 |
|  |  | Weighted median | 46 | 0.25 | 1.12 | 0.92 | 1.36 |
|  |  | Inverse variance weighted | 46 | 0.02 | 1.23 | 1.03 | 1.47 |
|  |  | Simple mode | 46 | 0.43 | 1.16 | 0.80 | 1.69 |
|  |  | Weighted mode | 46 | 0.77 | 1.03 | 0.84 | 1.27 |
|  | Lacunar stroke | MR Egger | 46 | 0.17 | 0.60 | 0.29 | 1.22 |
|  |  | Weighted median | 46 | 0.11 | 0.69 | 0.44 | 1.08 |
|  |  | Inverse variance weighted | 46 | 0.98 | 1.00 | 0.71 | 1.43 |
|  |  | Simple mode | 46 | 0.75 | 0.87 | 0.36 | 2.09 |
|  |  | Weighted mode | 46 | 0.10 | 0.64 | 0.38 | 1.08 |
| FI | Ischemic stroke | MR Egger | 14 | 0.88 | 1.17 | 0.17 | 7.77 |
|  |  | Weighted median | 14 | 0.02 | 1.66 | 1.07 | 2.58 |
|  |  | Inverse variance weighted | 14 | 0.02 | 1.46 | 1.05 | 2.02 |
|  |  | Simple mode | 14 | 0.11 | 1.81 | 0.91 | 3.58 |
|  |  | Weighted mode | 14 | 0.14 | 1.83 | 0.87 | 3.88 |
|  | Lacunar stroke | MR Egger | 14 | 0.42 | 6.19 | 0.08 | 450.69 |
|  |  | Weighted median | 14 | 0.37 | 1.62 | 0.57 | 4.62 |
|  |  | Inverse variance weighted | 14 | 0.28 | 1.53 | 0.71 | 3.32 |
|  |  | Simple mode | 14 | 0.44 | 2.07 | 0.35 | 12.30 |
|  |  | Weighted mode | 14 | 0.69 | 0.69 | 0.11 | 4.18 |
| 2h-glu | Ischemic stroke | MR Egger | 17 | 0.81 | 0.96 | 0.69 | 1.34 |
|  |  | Weighted median | 17 | 0.27 | 1.07 | 0.95 | 1.20 |
|  |  | Inverse variance weighted | 17 | 0.05 | 1.12 | 1.00 | 1.25 |
|  |  | Simple mode | 17 | 0.67 | 1.05 | 0.86 | 1.28 |
|  |  | Weighted mode | 17 | 0.91 | 1.01 | 0.86 | 1.18 |
|  | Lacunar stroke | MR Egger | 15 | 0.10 | 1.62 | 0.96 | 2.75 |
|  |  | Weighted median | 15 | 0.03 | 1.32 | 1.02 | 1.70 |
|  |  | Inverse variance weighted | 15 | 0.01 | 1.26 | 1.05 | 1.51 |
|  |  | Simple mode | 15 | 0.87 | 1.04 | 0.63 | 1.72 |
|  |  | Weighted mode | 15 | 0.07 | 1.53 | 1.00 | 2.34 |

***Abbreviations:*** *n: number; HbA1c: Hemoglobin A1c; FG: Fasting glucose; FI: Fasting insulin; 2h-glu: 2-hour glucose; SNP: Single nucleotide polymorphism; OR: Odds ratio; LCI: Lower confidence interval; UCI: Upper confidence interval.*

**Supplementary Table S7. Mendelian randomization analysis for the causal associations between T2D and brain MRI markers based on five methods**

| Outcomes | Method | nSNP | p-value | OR | LCI | UCI |
| --- | --- | --- | --- | --- | --- | --- |
| Brain volume | MR Egger | 405 | 0.91 | 1.00 | 0.95 | 1.05 |
|  | Weighted median | 405 | 0.83 | 1.00 | 0.96 | 1.04 |
|  | Inverse variance weighted | 405 | 0.47 | 0.99 | 0.97 | 1.01 |
|  | Simple mode | 405 | 0.95 | 1.00 | 0.91 | 1.09 |
|  | Weighted mode | 405 | 0.73 | 0.99 | 0.95 | 1.04 |
| Grey matter volume | MR Egger | 405 | 0.49 | 1.02 | 0.97 | 1.07 |
|  | Weighted median | 405 | 0.70 | 0.99 | 0.96 | 1.03 |
|  | Inverse variance weighted | 405 | 0.00 | 0.97 | 0.95 | 0.99 |
|  | Simple mode | 405 | 0.14 | 0.93 | 0.84 | 1.02 |
|  | Weighted mode | 405 | 0.37 | 1.02 | 0.97 | 1.07 |
| White matter volume | MR Egger | 405 | 0.45 | 0.98 | 0.93 | 1.03 |
|  | Weighted median | 405 | 0.47 | 0.99 | 0.96 | 1.02 |
|  | Inverse variance weighted | 405 | 0.21 | 1.02 | 0.99 | 1.04 |
|  | Simple mode | 405 | 0.17 | 1.07 | 0.97 | 1.18 |
|  | Weighted mode | 405 | 0.17 | 0.97 | 0.93 | 1.01 |
| Hippocampus volume | MR Egger | 405 | 0.59 | 0.96 | 0.85 | 1.10 |
|  | Weighted median | 405 | 0.44 | 0.96 | 0.86 | 1.07 |
|  | Inverse variance weighted | 405 | 0.18 | 1.04 | 0.98 | 1.11 |
|  | Simple mode | 405 | 0.89 | 0.98 | 0.75 | 1.29 |
|  | Weighted mode | 405 | 0.41 | 0.94 | 0.81 | 1.09 |
| WMH | MR Egger | 405 | 0.71 | 0.99 | 0.94 | 1.04 |
|  | Weighted median | 405 | 0.60 | 0.99 | 0.95 | 1.03 |
|  | Inverse variance weighted | 405 | 0.83 | 1.00 | 0.98 | 1.03 |
|  | Simple mode | 405 | 0.92 | 1.00 | 0.92 | 1.09 |
|  | Weighted mode | 405 | 0.50 | 0.98 | 0.94 | 1.03 |

***Abbreviations:*** *n: number; T2D: Type 2 diabetes; WMH: White matter hyperintensities; SNP: Single-nucleotide polymorphism; OR: Odds ratio; LCI: Lower confidence interval; UCI: Upper confidence interval.*

**Supplementary Table S8. Mendelian randomization analysis for the causal associations between glycemic traits and brain MRI markers based on five methods**

| Exposures | Outcomes | Method | nSNP | p-value | OR | LCI | UCI |
| --- | --- | --- | --- | --- | --- | --- | --- |
| HbA1c | Brain volume | MR Egger | 174 | 0.77 | 1.04 | 0.82 | 1.32 |
|  |  | Weighted median | 174 | 0.47 | 0.94 | 0.80 | 1.11 |
|  |  | Inverse variance weighted | 174 | 0.06 | 0.89 | 0.78 | 1.00 |
|  |  | Simple mode | 174 | 0.51 | 0.87 | 0.59 | 1.30 |
|  |  | Weighted mode | 174 | 0.98 | 1.00 | 0.80 | 1.24 |
|  | Grey matter volume | MR Egger | 174 | 0.36 | 1.12 | 0.88 | 1.41 |
|  |  | Weighted median | 174 | 0.92 | 1.01 | 0.85 | 1.19 |
|  |  | Inverse variance weighted | 174 | 0.59 | 0.97 | 0.86 | 1.09 |
|  |  | Simple mode | 174 | 0.91 | 1.02 | 0.68 | 1.53 |
|  |  | Weighted mode | 174 | 0.82 | 1.02 | 0.83 | 1.26 |
|  | White matter volume | MR Egger | 174 | 0.73 | 0.96 | 0.75 | 1.23 |
|  |  | Weighted median | 174 | 0.29 | 0.91 | 0.76 | 1.08 |
|  |  | Inverse variance weighted | 174 | 0.02 | 0.85 | 0.75 | 0.97 |
|  |  | Simple mode | 174 | 0.22 | 0.75 | 0.47 | 1.19 |
|  |  | Weighted mode | 174 | 0.65 | 0.95 | 0.76 | 1.19 |
|  | Hippocampus volume | MR Egger | 170 | 0.89 | 0.96 | 0.54 | 1.71 |
|  |  | Weighted median | 170 | 0.28 | 0.75 | 0.45 | 1.26 |
|  |  | Inverse variance weighted | 170 | 0.00 | 0.61 | 0.45 | 0.83 |
|  |  | Simple mode | 170 | 0.43 | 0.65 | 0.22 | 1.91 |
|  |  | Weighted mode | 170 | 0.30 | 0.72 | 0.38 | 1.34 |
|  | WMH | MR Egger | 174 | 0.11 | 1.21 | 0.96 | 1.52 |
|  |  | Weighted median | 174 | 0.23 | 1.11 | 0.93 | 1.33 |
|  |  | Inverse variance weighted | 174 | 0.90 | 1.01 | 0.89 | 1.14 |
|  |  | Simple mode | 174 | 0.44 | 1.17 | 0.79 | 1.72 |
|  |  | Weighted mode | 174 | 0.36 | 1.11 | 0.89 | 1.37 |
| FG | Brain volume | MR Egger | 48 | 0.98 | 1.00 | 0.75 | 1.32 |
|  |  | Weighted median | 48 | 0.90 | 0.99 | 0.84 | 1.17 |
|  |  | Inverse variance weighted | 48 | 0.12 | 0.90 | 0.78 | 1.03 |
|  |  | Simple mode | 48 | 0.32 | 1.21 | 0.84 | 1.75 |
|  |  | Weighted mode | 48 | 0.92 | 0.99 | 0.82 | 1.19 |
|  | Grey matter volume | MR Egger | 48 | 0.44 | 0.89 | 0.66 | 1.20 |
|  |  | Weighted median | 48 | 0.44 | 0.93 | 0.77 | 1.12 |
|  |  | Inverse variance weighted | 48 | 0.13 | 0.89 | 0.77 | 1.03 |
|  |  | Simple mode | 48 | 0.64 | 1.10 | 0.75 | 1.61 |
|  |  | Weighted mode | 48 | 0.65 | 0.95 | 0.78 | 1.17 |
|  | White matter volume | MR Egger | 48 | 0.42 | 1.10 | 0.87 | 1.40 |
|  |  | Weighted median | 48 | 0.45 | 1.06 | 0.91 | 1.24 |
|  |  | Inverse variance weighted | 48 | 0.25 | 0.93 | 0.83 | 1.05 |
|  |  | Simple mode | 48 | 1.00 | 1.00 | 0.72 | 1.38 |
|  |  | Weighted mode | 48 | 0.35 | 1.08 | 0.92 | 1.28 |
|  | Hippocampus volume | MR Egger | 48 | 0.30 | 0.71 | 0.37 | 1.35 |
|  |  | Weighted median | 48 | 0.30 | 0.77 | 0.47 | 1.26 |
|  |  | Inverse variance weighted | 48 | 0.32 | 0.85 | 0.62 | 1.17 |
|  |  | Simple mode | 48 | 0.76 | 1.15 | 0.45 | 2.93 |
|  |  | Weighted mode | 48 | 0.54 | 0.84 | 0.47 | 1.47 |
|  | WMH | MR Egger | 48 | 0.35 | 1.16 | 0.85 | 1.58 |
|  |  | Weighted median | 48 | 0.41 | 0.93 | 0.77 | 1.11 |
|  |  | Inverse variance weighted | 48 | 0.45 | 0.94 | 0.81 | 1.10 |
|  |  | Simple mode | 48 | 0.07 | 0.68 | 0.46 | 1.02 |
|  |  | Weighted mode | 48 | 0.39 | 0.90 | 0.71 | 1.14 |
| FI | Brain volume | MR Egger | 14 | 0.60 | 1.56 | 0.31 | 7.84 |
|  |  | Weighted median | 14 | 0.96 | 1.01 | 0.68 | 1.51 |
|  |  | Inverse variance weighted | 14 | 0.48 | 0.90 | 0.67 | 1.21 |
|  |  | Simple mode | 14 | 0.69 | 1.15 | 0.58 | 2.29 |
|  |  | Weighted mode | 14 | 0.66 | 1.17 | 0.60 | 2.30 |
|  | Grey matter volume | MR Egger | 14 | 0.24 | 3.36 | 0.50 | 22.71 |
|  |  | Weighted median | 14 | 0.47 | 0.86 | 0.56 | 1.31 |
|  |  | Inverse variance weighted | 14 | 0.18 | 0.78 | 0.54 | 1.12 |
|  |  | Simple mode | 14 | 0.95 | 1.03 | 0.44 | 2.39 |
|  |  | Weighted mode | 14 | 0.94 | 1.03 | 0.52 | 2.04 |
|  | White matter volume | MR Egger | 14 | 0.77 | 0.78 | 0.16 | 3.93 |
|  |  | Weighted median | 14 | 0.45 | 1.16 | 0.79 | 1.71 |
|  |  | Inverse variance weighted | 14 | 0.63 | 1.07 | 0.80 | 1.44 |
|  |  | Simple mode | 14 | 0.43 | 1.33 | 0.66 | 2.67 |
|  |  | Weighted mode | 14 | 0.59 | 1.22 | 0.60 | 2.52 |
|  | Hippocampus volume | MR Egger | 14 | 0.44 | 12.88 | 0.03 | 6,566.09 |
|  |  | Weighted median | 14 | 0.80 | 1.18 | 0.32 | 4.32 |
|  |  | Inverse variance weighted | 14 | 0.41 | 1.59 | 0.53 | 4.80 |
|  |  | Simple mode | 14 | 0.76 | 0.69 | 0.07 | 7.30 |
|  |  | Weighted mode | 14 | 0.84 | 0.80 | 0.10 | 6.67 |
|  | WMH | MR Egger | 14 | 0.41 | 2.16 | 0.36 | 12.94 |
|  |  | Weighted median | 14 | 0.39 | 1.21 | 0.79 | 1.85 |
|  |  | Inverse variance weighted | 14 | 0.08 | 1.33 | 0.97 | 1.82 |
|  |  | Simple mode | 14 | 0.85 | 0.93 | 0.43 | 2.02 |
|  |  | Weighted mode | 14 | 0.89 | 0.94 | 0.41 | 2.14 |
| 2h-glu | Brain volume | MR Egger | 18 | 0.29 | 0.91 | 0.76 | 1.08 |
|  |  | Weighted median | 18 | 0.05 | 0.92 | 0.84 | 1.00 |
|  |  | Inverse variance weighted | 18 | 0.05 | 0.94 | 0.88 | 1.00 |
|  |  | Simple mode | 18 | 0.59 | 0.96 | 0.84 | 1.10 |
|  |  | Weighted mode | 18 | 0.18 | 0.92 | 0.82 | 1.03 |
|  | Grey matter volume | MR Egger | 18 | 0.91 | 0.99 | 0.79 | 1.23 |
|  |  | Weighted median | 18 | 0.43 | 0.96 | 0.88 | 1.06 |
|  |  | Inverse variance weighted | 18 | 0.08 | 0.93 | 0.87 | 1.01 |
|  |  | Simple mode | 18 | 0.53 | 0.95 | 0.80 | 1.12 |
|  |  | Weighted mode | 18 | 0.47 | 0.95 | 0.82 | 1.10 |
|  | White matter volume | MR Egger | 18 | 0.20 | 0.88 | 0.72 | 1.07 |
|  |  | Weighted median | 18 | 0.59 | 0.97 | 0.88 | 1.08 |
|  |  | Inverse variance weighted | 18 | 0.25 | 0.96 | 0.90 | 1.03 |
|  |  | Simple mode | 18 | 0.16 | 0.88 | 0.75 | 1.04 |
|  |  | Weighted mode | 18 | 0.08 | 0.87 | 0.75 | 1.01 |
|  | Hippocampus volume | MR Egger | 18 | 0.38 | 0.76 | 0.42 | 1.38 |
|  |  | Weighted median | 18 | 0.11 | 0.79 | 0.59 | 1.05 |
|  |  | Inverse variance weighted | 18 | 0.12 | 0.85 | 0.69 | 1.05 |
|  |  | Simple mode | 18 | 0.28 | 0.76 | 0.47 | 1.23 |
|  |  | Weighted mode | 18 | 0.26 | 0.78 | 0.52 | 1.18 |
|  | WMH | MR Egger | 18 | 0.47 | 0.91 | 0.70 | 1.17 |
|  |  | Weighted median | 18 | 0.02 | 1.12 | 1.02 | 1.24 |
|  |  | Inverse variance weighted | 18 | 0.14 | 1.07 | 0.98 | 1.18 |
|  |  | Simple mode | 18 | 0.34 | 1.09 | 0.92 | 1.31 |
|  |  | Weighted mode | 18 | 0.13 | 1.12 | 0.97 | 1.29 |

***Abbreviations:*** *n: number; HbA1c: Hemoglobin A1c; FG: Fasting glucose; FI: Fasting insulin; 2h-glu: 2-hour glucose; WMH: White matter hyperintensities; SNP: Single-nucleotide polymorphism; OR: Odds ratio; LCI: Lower confidence interval; UCI: Upper confidence interval.*
